# Genosolver: Rare Disease Diagnosis through Holistic Integration of Unstructured Clinical Narratives Using Large Language and Reasoning Models

**DOI:** 10.64898/2026.06.04.26354845

**Authors:** Tanhim Islam, Martin Danner, Zain Ziad, Matthias Begemann, Danique Beijer, Annette Lischka, Eva Lausberg, Larissa Mattern, Julia Suh, Pauline Wittig, Nergis Güzel, Elia Schlaich, Radina Karaivanova, Sofia D’Augello, Lena Franken, Jarik Rüdebusch, Robin Müller, Eva Perchalla, Hans Zempel, Natja Haag, Katja Eggermann, Thomas Eggermann, Robert Meyer, Florian Kraft, Miriam Elbracht, Ingo Kurth, Jeremias Krause

## Abstract

**Background:** Molecular medicine has made genetic diagnostics crucial for rare diseases, but the majority of patients remains without diagnosis even after state-of-the-art assessment. Standardized systems for integrating clinical features, such as the Human Phenotype Ontology (HPO), offer assistance, but are often insufficiently detailed and fail to capture crucial clinical parameters such as age at onset, longitudinal changes in symptoms, detailed characteristics of a clinical symptom, or the absence of a feature.

**Results:** We present Genosolver an integrated workflow that utilizes machine learning to address this bottleneck. Using Large Language Models (LLMs) and Large Reasoning Models (LRMs) on unstructured clinical notes and electronic health care data, we generate a workflow that unifies phenotype extraction, generates differential diagnosis, and prioritizes genetic variants from genome data. We evaluated the performance on 233 previously genetically solved cases, where Genosolver ranked the causative gene first in 72% of cases and in 94% of cases in the top 10 gene list, outperforming the existing benchmarking tool Exomiser by 9%. Semi-automated reanalysis of 1,875 unsolved rare disease cases yielded an additional diagnostic rate of 1.7%. Incorporating rich, unstandardized clinical narratives substantially enhanced model performance beyond HPO-only inputs and demonstrated competitive results using data security compliant local models.

**Conclusion:** Integrating unstandardized clinical data with local LLMs and reasoning offers a scalable, data-secure workflow that increases molecular diagnoses in rare diseases.

## 1. Background

Molecular medicine has profoundly changed patient care, establishing genetic profiles as fundamental guides in rare diseases and oncology^1–7^. While sequencing technologies have advanced, this progress has created a significant diagnostic bottleneck: interpreting the vast amounts of genomic data generated by whole-genome or whole-exome sequencing (WGS/WES)^8,9^. Computational approaches are utilized to annotate and prioritize variants using multiple data modalities—including sequence information, population frequencies, and standardized clinical features^10–28^.

The encoding of clinical phenotypes to aid diagnosis is typically managed by systems like the Human Phenotype Ontology (HPO), which provides invaluable standardization^29,30^. However, the HPO has inherent limitations in capturing complex clinical information, such as temporal and longitudinal symptom changes, detailed family history, or negated findings. While some advanced machine learning models exist to integrate additional and more continuous data (e.g., from photographs)^31^, current prioritization algorithms remain fragmented.

Recent advances using Large Language Models (LLMs) have shown promise for rare disease diagnosis by enabling phenotype extraction and differential diagnosis^32–35^. However, existing implementations often fail to bridge the entire pipeline. They are typically limited either to converting features into HPO terms, thus losing crucial natural language details, or they integrate genetic information only superficially. To our knowledge, there is currently no open-source pipeline that integrates all three steps: phenotype extraction, rare disease diagnosis, and variant prioritization into a single workflow capable of working with both standardized systems like the HPO and non-standardized human phenotypic features.

To address this unmet need, we have implemented Genosolver, a multi-stage workflow based on state-of-the-art LLMs and Large Reasoning Models (LRMs). Genosolver is designed to work seamlessly with both standardised HPO terms and extensive, non-standardised clinical descriptions, thereby providing a unified, integrated platform for complex diagnostic workflows in rare diseases.

## 2. Results

### Genosolver: A Large Language Model Workflow for Phenotype Extraction, Rare Disease Diagnosis and Phenotypical Variant Prioritization

Genosolver is a machine learning (ML) workflow designed for phenotype extraction, rare disease diagnosis, and variant prioritization from high-throughput sequencing data. An overview of the workflow is shown in **Figure 1**. An in-depth analysis and description of the workflow is provided as supplementary chapter. Genosolver consists of three modules:

1. Phenotype extractor: Extracts phenotypic features from unstructured electronic health records (EHR). This module was benchmarked by using a by-proxy evaluation, by testing how well Genosolver can extract HPO terms.
2. Disease designator: Identifies potential diagnoses by searching for a vector database, a specialized data structure that enables ehicient similarity searches containing phenotypic descriptions, their synonyms, and associated genes. This module is used as a coarse phenotype filter.
3. Variant prioritizor: Matches the potential diagnoses with a filtered list of the patient’s genetic variants. This step uses LLMs and LRMs with a retrieval-augmented-generation (RAG)^34^ approach, which incorporates curated literature evidence retrieved from a vector store. The matched variants and diseases are then evaluated to determine whether they explain the patient’s phenotype.

**Figure 1:**
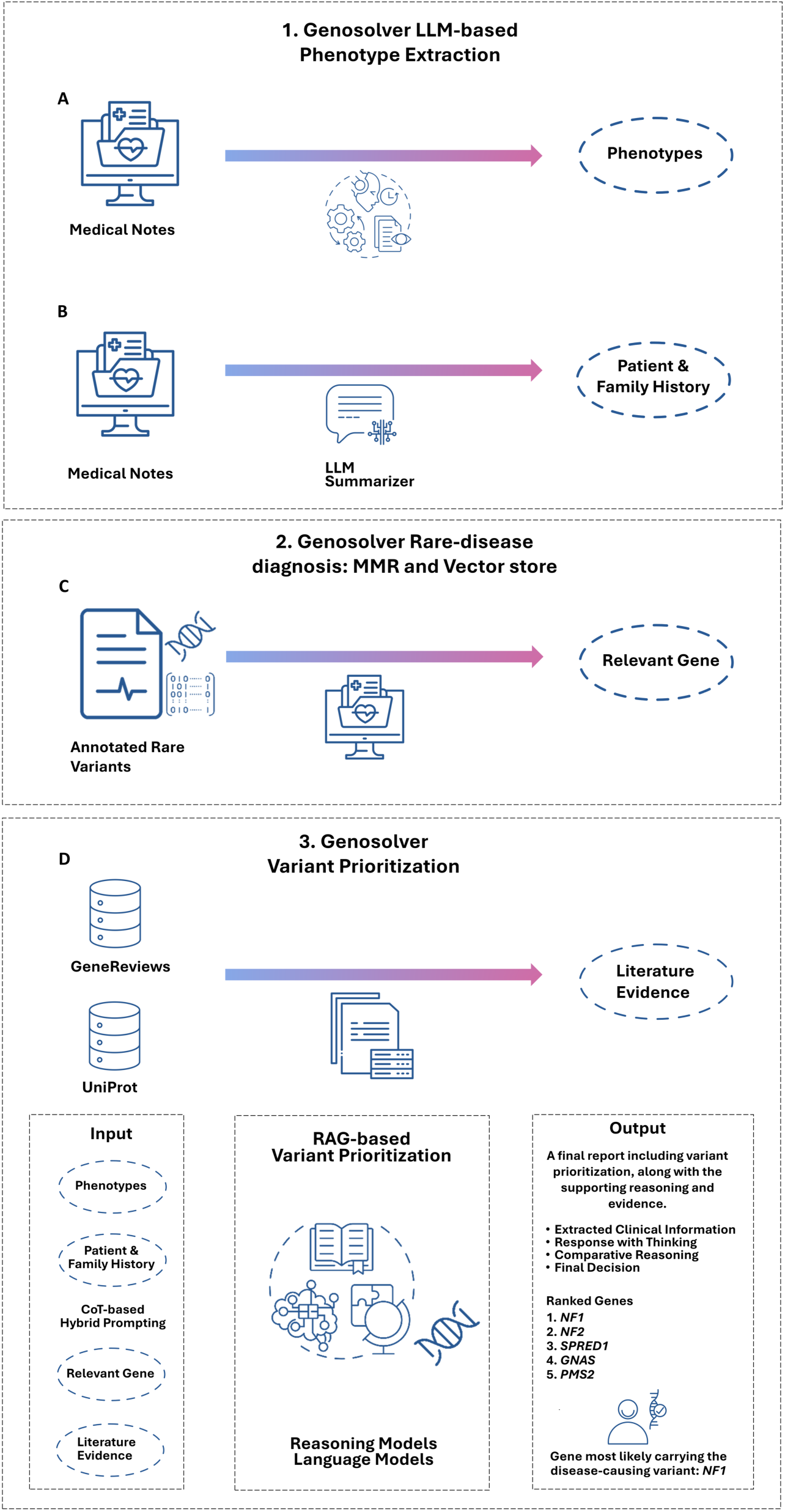
Overview of Genosolver: 1) At first, we evaluated how well LLMs can be utilized to extract standardized and non-standardized phenotypic features and information regarding clinical context from structured and unstructured EHRs. 2) Subsequentially, the extracted phenotypic features are utilized to query a vector store, to find potentially fitting rare-disease-gene associations. 3) Finally, the phenotypic features are matched with a filtered list of the genetic variants of the patient and passed to LLMs and LRMs (combined with a RAG approach), to evaluate whether the gene containing the disease-causing variant has been found, which is done by ranking the input variants. A reasoning summary is created, explaining the ranking process.

### Variant Prioritization using DiEerent Large Language Models

We explored the capabilities of Genosolver to perform phenotypic driven genetic variant prioritization, by evaluating it on a cohort of 233 solved cases with rare diseases. The diseases were classified into four categories which were neurodevelopmental disorders (n=102), organ-specific diseases (n=51) (such as heart, kidney, liver), neuromuscular disorders (n=49), and hematological disorders and neoplasms (n=31) (**Supplementary Figure 1**).105 patients of the study were male, 128 patients were female, the median age of the group was 25.5 years. For variant prioritization we explored four different LLMs which are usable in a data secure local set up. These models were: Llama-3.3-70B-Instruct^36^, MedGo 32B based on the Qwen3 architecture^37^, DeepSeek^38,39^ distilled Llama 70B, and GPT OSS 120B^40^. The evaluation was performed using the hit rate at k (i.e. the correct gene was identified) with top 1, top 3, top 5 and top 10 thresholds (“recall@k”; k=1; k=3; k=5; k=10). As a baseline model we selected Exomiser^41^, a renowned variant prioritization tool, capable of ranking genetic variants based on phenotypic and molecular features. In total we performed the following experiments, each with differently prepared phenotypical information as an input:

1. Unstandardized Phenotypes and Clinical Context: Utilizing unstandardized phenotypic features and automatically extracted information relevant for family and clinical history.
2. Expert assigned HPOs: Utilizing expert assigned HPO-terms.
3. Machine generated HPOs: Utilizing HPO-terms generated by the PhenoBERT^42^ Llama^36^ fusion, as detailed in the phenotype extraction supplementary chapter.
4. Fusing Genosolver with Exomisers automated ACMG^43^-classification, by filtering out Variants classified as benign or likely-benign by Exomiser.
5. Fusing Genosolver with Exomisers automated ACMG^43^-classification and Gene-Variant score, by filtering out Variants classified as benign or likely-benign by Exomiser, as well as keeping only variants with a high variant prioritization score. In addition, utilizing both phenotypic features and expert-assigned HPO-terms.

The results of the best performing model versus Exomiser across the Experiments are given in **Figure 2**, a comparison between all models can be seen in **Supplementary Figure 2**. Among the three head-to-head comparisons between Exomiser and Genosolver the best performing model was the Genosolver model from experiment 1, which uses unstructured clinical notes, incorporating the GPT OSS 120B model. It reached hit rates of 0.72 (k=1), 0.83 (k=3), 0.89 (k=5), and 0.94 (k=10) (**Figure 2**). The second-best approach was the Exomiser total score achieving hit rates of 0.63 (k=1), 0.78 (k=3), 0.81 (k=5) and 0.85 (k=10). The other three LLMs (Llama, MedGo and DeepSeek distilled) alone failed to surpass the baseline across all three experiments. Comparing Genosolver GPT OSS 120B to GPT 5.1 with different reasoning settings highlighted comparable values regarding recall at 5 and 10 (Supplementary chapter: Comparison to closed weight models).

**Figure 2:**
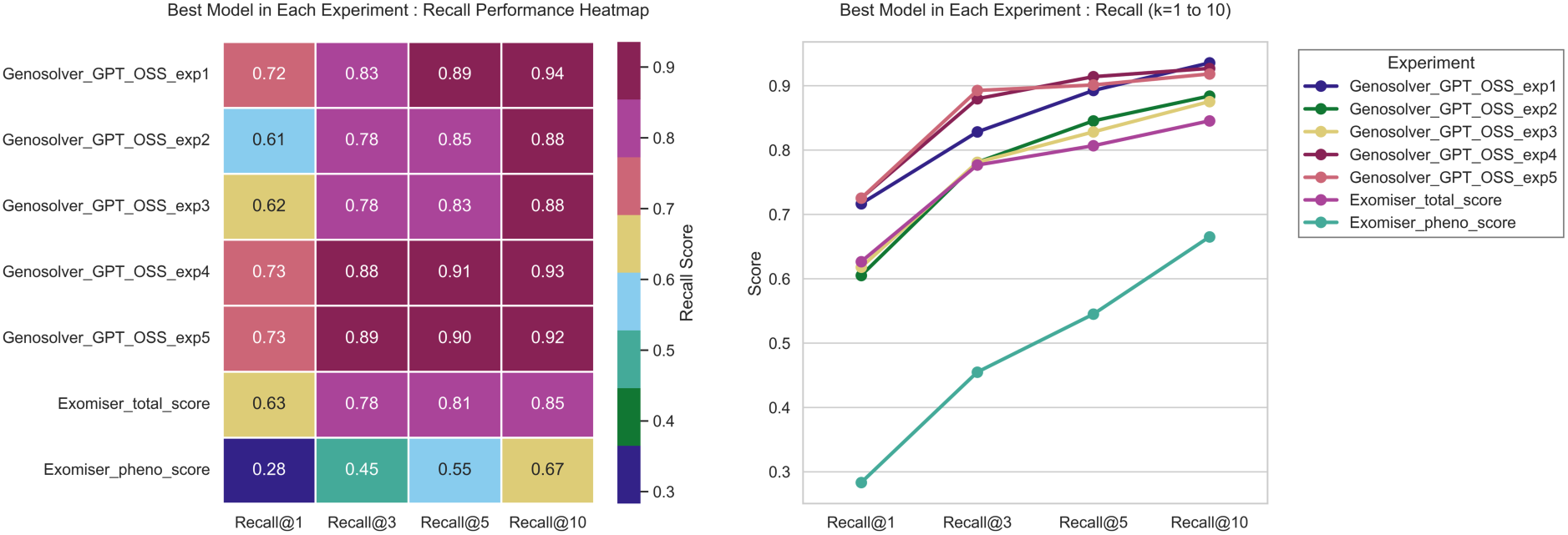
Head-to-head comparison between Genosolver and Exomiser for the best performing model. A) Heatmap visualization showing the Recall Score achieved by each best performing model (per experiment) across different recall thresholds (k=1,3,5,10). K=1 means, that the correct gene was ranked as number 1, k=3 means, that the correct gene was within the first three genes, etc. B) Line graph detailing the best preforming model, in comparison to the benchmark Exomiser (total score) and the Exomiser pheno-score. Across all thresholds, there is at least one version of Genosolver that outperforms the Exomiser.

In both experiment 2 and 3, the performance of the GPT OSS 120B model also dropped below the Exomiser^41^ performance except for the hit rate of 0.86 (k=10) which it achieved in both experiments.

The fusion strategy in experiment 4 enhanced Genosolver’s performance, independently of the incorporated model. The Genosolver workflow integrating the GPT OSS 120B model stayed the best approach and achieved hit rates of 0.73 (k=1), 0.88 (k=3), 0.91 (k=5) and 0.93 (k=10).

Utilizing the strategy planned for the experiment 5 resulted in mixed results. While all models still outperformed the baseline, a shift towards a decreasing performance was visible at recall at 5 and 10. Here the best results were achieved by the Genosolver workflow which integrated the GPT OSS model with hit rates of 0.72 (k=1), 0.85 (k=3), 0.88 (k=5) and 0.91 (k=10). The performance of the workflow integrating the GPT OSS model dropped noticeably to hit rates of 0.65 (k=1), 0.81 (k=3), 0.82 (k=5) and 0.83 (k=10).

### Comparing Unstructured Medical Phenotypic Features with HPO Terms

To further explore the additional value of non-standardized phenotypic features compared with defined HPO terms, we mapped the unstandardized phenotypic features to the expert assigned HPO terms on a case-by-case basis to identify unique unmapped terms. In total we found 522 phenotype terms which were extracted by the Genosolver workflow, and which could (either partially or fully) not be mapped to assigned or existing HPO terms. These included: (1) phenotypical features missed by experts (n=342); (2) specific phenotypes lacking corresponding HPO terms (n=34); (3) additional attributes not captured in assigned HPOs (n=131), such as "therapy-resistant myoclonic-astatic epilepsy"; (4) time components within descriptions (n=67) ("first febrile convulsion at about 5 years"); (5) multi-level phenotypic chains (n=44), such as “heart defect with enlarged right ventricle and mild atrial enlargement”; (6) therapeutic agents not captured by the current HPO (n=34); (7) diagnostic results not captured by the current HPO (n=34), e.g., "EEG normal" or "SDHB-deficient paraganglioma"; (8) Negative features (n=8) like “absent EEG findings”; (9) family history with relevant phenotypes (n=6) ("night blindness in the father"). Since the Genosolver workflow also ingests information on sex, age and family history, we wanted to explore how many times the model picks up on this information in its input and utilizes them in its formulated argumentation. For this we utilized the overall best performing model GPT OSS 120B. This model utilized family information (e.g. a plausible inheritance pattern) in its argumentation in 48 cases, age in 43 cases and sex in 9 cases. We subsequently evaluated the correctness of the argumentation revealing that the family information and inheritance pattern was explained correct in 47/48 cases. The criteria for age were applied correctly in all cases, where age was mentioned. In 6 cases the model utilized the patients’ sex correctly to justify a disease as more plausible (e.g. visible in figure 4B), however, in 3 cases it provided an ambiguous answer. In all these three cases the model argued that the sex aligns well with the present diagnosis for this patient, however in each of these 3 cases the diagnosis was for a disease for which no obvious sex bias was known, while for some of the present differential diagnoses a sex bias was known.

**Figure 4:**
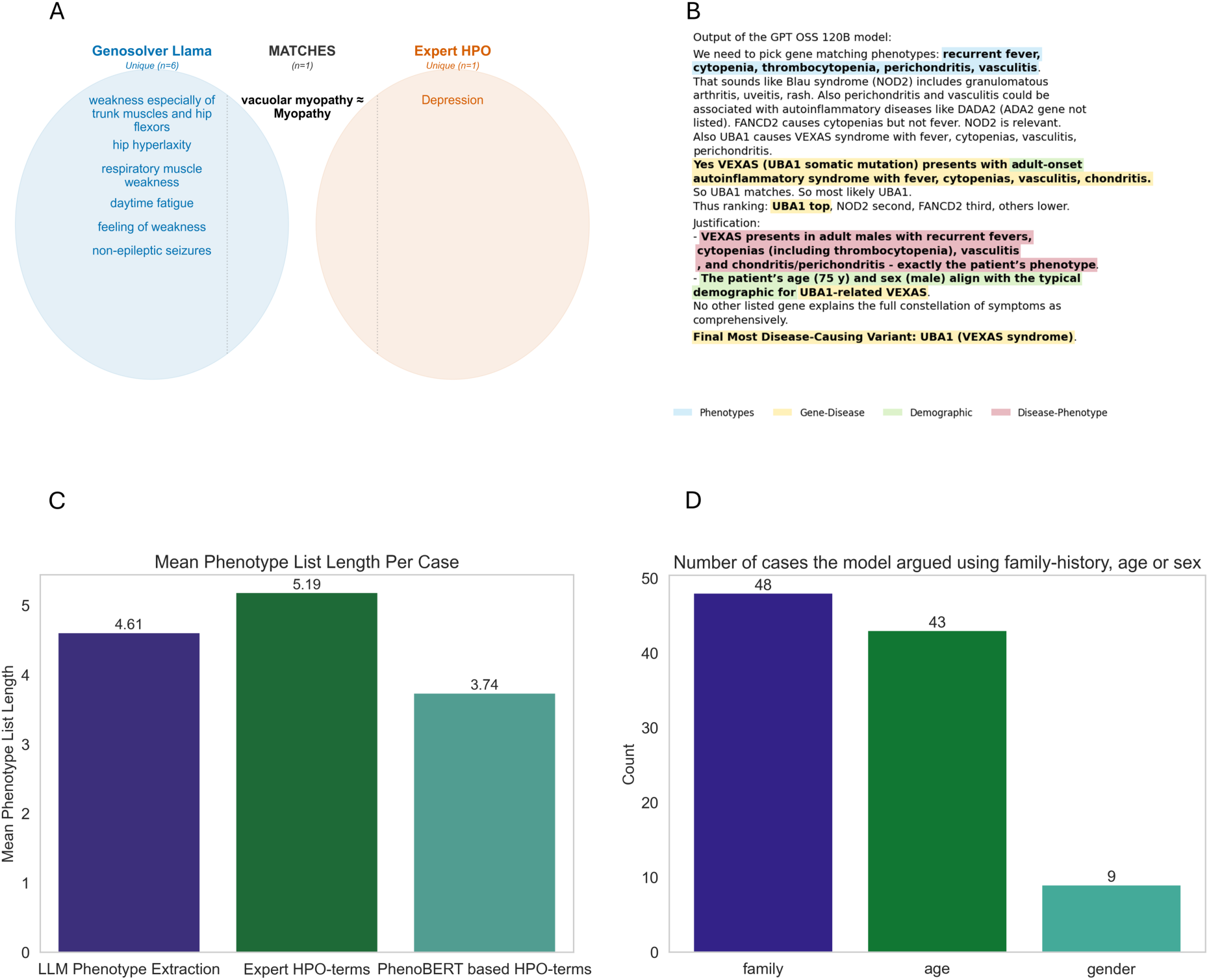
A) This Venn diagram compares the phenotype extraction capabilities of Genosolver LLM against expert-defined HPO terms, using the same patient’s medical notes for both. Match was done via fuzzy matching through difflib SequenceMatcher, followed by a greedy algorithm prioritizing highest-score pairs for selection. B) This figure illustrates the reasoning correctness of the GPT OSS model, demonstrating its ability to correctly identify the most probable genes based on phenotypic features and sex, leading to correct diagnoses. A more in-depth analysis of the output of Genosolver for this case can be found in the supplementary material (including supplementary figure 1) C) This bar graph presents the average number of phenotypic terms generated per case across all conducted experiments. D) This bar graph quantifies the number of cases that incorporated family history, age, and sex information, using both Genosolver LLM phenotype extraction and the LLM-summarized medical notes.

### Performing a Semi-Automized Large Scale Reanalysis using Genosolver

Finally, we wanted to evaluate whether LLM-based phenotypic prioritization can also be usefully utilized in large scale semi-automized reanalysis. For this we performed two evaluations:

1. We repeated the previously presented variant prioritization experiment with the same 233 previously solved cases, with the newest Exomiser version (15.0.0)^44^ in conjunction with the recently published 2512 dataset version for Exomiser^44^. This ensured that the majority of variants in the dataset were annotated as pathogenic or likely pathogenic (as the 2,512 dataset contains the updated ClinVar annotation for these cases) and that Exomiser^41^ had access to these recent annotations, enabling an evaluation by proxy of what might have been detected if these annotations had been available at the time of the original analysis. These results are visualized in **Figure 5**.
2. Subsequentially, we utilized the Genosolver to perform a large-scale reanalysis on a cohort (n=1,875) of previously unsolved cases that underwent exome-sequencing.

**Figure 5:**
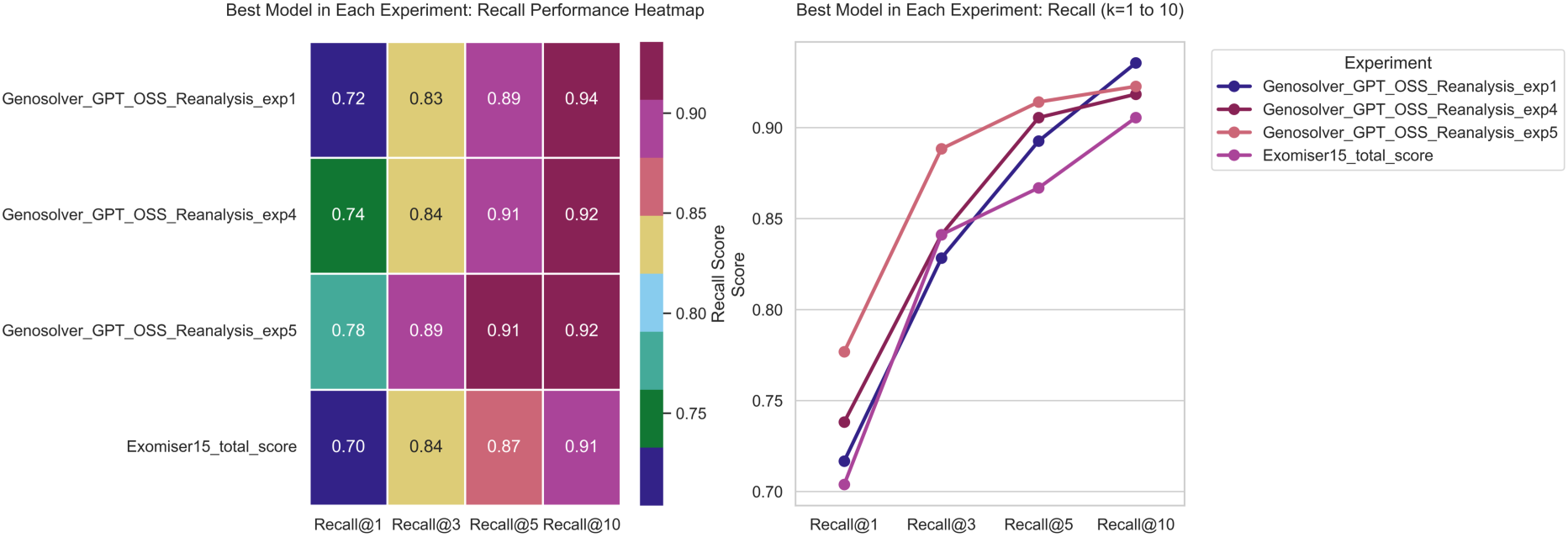
Head-to-head comparison between Genosolver and Exomiser for the best performing model. A) Heatmap visualization showing the Recall Score achieved by each best performing model (per experiment) across di[erent recall thresholds (k=1,3,5,10B) Line graph detailing the best preforming model, in comparison to the benchmark Exomiser (total score).. These results were obtained with the Exomiser 15.0.0 version and the 2512 dataset version.

The Exomiser (15.0.0)^44^ achieved hit rates of 0.70 (k=1), 0.84 (k=3), 0.87 (k=5) and 0.91 (k=10); thereby underperforming compared to the hit rates (k=1,5,10) of the standalone Genosolver workflow which integrates the GPT OSS 120B model, although outperforming the standalone Genosolver workflow (integrating GPT OSS 120B model) at k=3. The other standalone versions of Genosolver (integrating Llama, MedGo and Deepseek distilled) were outperformed by that Exomiser (15.0.0) version. Additionally, similarly to the previous experiment, we also evaluated GPT OSS 120B standalone Genosolver and Exomiser (15.0.0)^44^ for a synergistic effect by evaluating the fusion approach of these two. Here we observed results comparable to the previous experiment. The best hit rates were achieved by the Genosolver workflow that integrated the GPT OSS 120B model. It achieved hit rates of 0.74 (k=1), 0.84 (k=3), 0.91 (k=5) and 0.92 (k=10). The remaining Genosolver versions (integrating Llama, MedGo and Deepseek distilled) again underperformed, compared to the standalone Exomiser (15.0.0.) version. Finally, we performed an additional fusion experiment. We observed that by utilizing the 2512 annotation data for Exomiser^44^ the number of ranked variants being associated with an OMIM annotation noticeably increased, leading to an increase of task complexity for the model. To evaluate whether reducing this added complexity would benefit the Genosolver models, we additionally utilized the Exomisers^44^ gene variant score. This score reflects a broad selection of features (pathogenicity predictions, allele frequency in the general population, constraint metrics for the genes etc.). Similarly to the utilisation of the Exomisers^44^ ACMG^43^ classification we utilized the Gene variant score to filter the gene input list which was subsequentially passed to the Genosolver variant prioritization framework. Utilizing this threshold fusion approach substantially improved the Genosolver hit rates. The overall best performing model was again the workflow integrating the GPT OSS 120 B model. It achieved hit rates of 0.78 (k=1), 0.89 (k=3), 0.91 (k=5) and 0.92 (k=10). The results of all experiments are visualized in **Supplementary figure 3**.

Having demonstrated a potential benefit for the utilization of LLMs in the reanalysis of previously unsolved cases we proceeded with testing the Genosolver workflow on an in-house cohort of 1,875 of patients which previously underwent exome analysis in suspicion of a rare disease. We chose cases from 2023 and 2024, allowing for a 2–3-year time gap between the previous diagnostics and this reanalysis project, which has been shown to be a reasonable time frame^45^. The cohort statistics are visualised in **Supplementary Figure 4**.

For this reanalysis we utilized the Llama model, since it was the most lightweight model, allowing a fast inference across the whole dataset. Additionally, the Exomiser^41^ output was filtered only for pathogenic or likely-pathogenic variants (as classified by Exomiser^41^ or assigned by ClinVar^46^). For newly added gene-variant annotations that were added after the first analysis, variants of uncertain clinical significance (VUS) were also kept. This shortened variant list was submitted to the Genosolver workflow, to find out whether any (new) diagnoses could be found. By following this approach, we found 31 new diagnoses corresponding to an additional diagnostic yield of 1.7%. This is in line with the diagnostic yield for SNVs and indels of other computational pipelines (e.g. Talos^45^). Subsequently, we evaluated the reasons for the change of outcome. In total three groups were formed: (1) change in ClinVar^46^ annotation (6 variants previously annotated as VUS); (2) change in gene annotation (2 variants involved genes which were previously without a gene-phenotype association); (3) prioritization strategy adjustment (21 variants discarded due to different prioritization strategies previously). In sum, this revisit-strategy enables genetic diagnosis of previously unsolved causes due changed annotation (both ClinVar^46^ or new gene), prioritization strategy setting adjustment, and in the majority of cases by integration of in-depth phenotypic information.

### Expanding Genosolver to additional variant types

Finally, we evaluated strategies to expand the Genosolver workflow to additional variant types. As a proof of concept we evaluated the following three additional experiments, utilizing additional cases, utilizing the Genosolver GPT OSS 120B fusion approach in:

1. Variant prioritization additionally including known repeat expansion disorders^47^. In brevity, for additional unrelated cases (n=5) diagnosed with a repeat expansion disorder, known repeat expansion disorders and their thresholds were extracted from the STRipy^48^ database. Subsequentially, ExpansionHunter^49^ was utilized and the resulting VCF files were filtered using the information extracted from STRipy^48^ and an in-house workflow. The filtered repeats were checked for phenotypical relevance using the Genosolver disease designator and followingly merged with the filtered indel and SNV list. This combined list was passed forward to the Genosolver variant prioritization. In this additional task Genosolver achieved a hit rate of 0.8 (k=1,3,5,10).
2. Variant prioritization additionally including single and multi-exon CNVs. Briefly summarized, for additional unrelated cases (n=10) with multi or single-exon CNVs, were processed with CNVkit^50^ and CNVizard^51^, as described before. The workflow previously presented in CNVizard^51^, was additionally refined to further reduce the amount of false positive calls, by only keeping single or multi-exon CNVs that were labelled as significant in CNVkits^50^ bintest, as well as having a log2-value above (amplifications) or below (deletions) the upper (amplifications) or lower (deletions) whisker in the log2-value boxplot of CNVkit^50^, while simultaneously being placed in the upper (amplifications) or lower (deletions) third of the whisker of the depth plot. The filtered CNVs were checked for phenotypical relevance using the Genosolver disease designator and followingly merged with the filtered indel and SNV list. This combined list was passed forward to the Genosolver variant prioritization. In this additional task Genosolver achieved hit rates of 0.8 (k=1) and 0.9 (k=3,5,10).
3. Phenotypic evaluation of multi-gene CNVs. For this Genosolver’s prioritization workflow had to be adapted. Instead of ranking individual genes and variants we adapted the Genosolver to enable the system to decide whether a CNV that contains multiple genes could be relevant for the patient’s phenotype or not. Again, CNVkit^50^ was utilized to identify consecutive CNVs involving multiple genes. Subsequentially, all genes involved in a CNV were consecutively passed to Genosolver, with the question whether this gene could be involved in the patient’s phenotype. If all genes were deemed to be irrelevant, the multi-gene CNV was discarded. Of the additional unrelated cases (n=9) with a multigene CNV Genosolver was correctly keeping the multi gene CNV in 7 out of 9 cases. The adjusted prompt utilized in this experiment can be found in supplementary table 8.

## 3. Discussion

### Phenotype Extraction and the Limits of the Current Version of the HPO

The HPO is the standard encoding system for rare diseases, offering a standardized and comprehensive characterisation of a large collection of phenotypic features which can be utilized by bioinformatic tools for variant prioritisation. Recently first evidence has been provided for the benefit of utilizing clinical information for rare disease diagnosis, which could not be encapsulated using the HPO^33^. They state that especially imaging findings, and clinical interventions are currently underrepresented in the HPO. These medical results can contain valuable information which can proof to be crucial for rare disease diagnosis. We expand on their findings, by transferring the concept to the field of genetic variant prioritization, enabling the Genosolver to operate with non-standardized phenotypic features. The performance gap between the experiment with the unstandardized phenotypic features and the experiment with the expert assigned HPO terms can partly be explained by a noticeable portion of missed HPO terms, which originally were present in the medical records. However, we argue that this cannot fully explain the performance gap between HPO terms and unstructured phenotypic features, since the workflow that utilized the HPO terms derived from the phenotypic features does not perform on par with the workflow working with unstandardized phenotypes. We provide evidence that, in line with the results of Yang, Huang and Lin et al.^33^, unstandardized phenotypic features can carry additional information (especially a more detailed description of phenotypic features) compared to what is captured by the HPO in its current state. We argue that this approach enables the use of LLMs as case summary tools, allowing natural language to serve as an interface for rare disease research and diagnostics, which has been recently proposed for the broader field of medicine^52^.

### Large Language Models for Variant Prioritization

Genosolver is a LLM based variant prioritization workflow that can be utilized to rank SNVs and indels by leveraging standardized and unstandardized phenotypic features. We demonstrate that the Genosolver workflow can achieve competitive performances while utilizing pretrained open-source models, such as Llama, MedGo, DeepSeek distilled and GPT OSS, allowing a privacy preserving local environment. An important difference, compared to available rare disease diagnosis workflows (such as RareSeek-R1^33^) is that the Genosolver workflow was evaluated using pretrained models, which were not further fine-tuned, but enhanced using a RAG-approach. This is crucial, since rare disease research is a rapidly evolving field^53^, meaning that each fine-tuned LLM is deemed to be outdated rather quickly; while a RAG-approach can be quickly updated to a new state of knowledge in this area. Furthermore, we evaluated the correctness and confidence of generated reports by Genosolver, highlighting that the model is capable to base its argumentations on additional available information such as the family history, inheritance patterns, age and sex.

### Integrating Large Language Models into reanalysis workflows

Reanalysing rare disease cases with updated annotation data has been shown to be effective previously^54^. We build onto this by additionally providing evidence that LLMs have potential to add additional benefit, by demonstrating that the most up to date version of Exomiser is outperformed by Genosolver (GPT OSS 120 B version). Additionally, among the newly solved 31 cases, 21 cases fall into the category of variants which were missed due to filter settings. This high proportion is in line with previous studies^45^, highlighting the limitations of rigid candidate gene list driven analysis, while simultaneously indicating a benefit of flexible phenotypic analysis, such as enabled by Genosolver.

## 4. Conclusion

The Genosolver workflow is an LLM-driven phenotypic variant prioritization tool that can operate independently or integrate data from Exomiser^41^. Demonstrated in both prospective and retrospective settings across large-scale reanalyses, this workflow can run locally within secure environments to import electronic health records, enhancing variant prioritization by combining LRMs with a RAG-based approach. The Genosolver can work with standardized and unstandardized phenotypical features. We demonstrated that unstandardized phenotypical features can carry information of additional clinical value. Finally, we provided a proof of concept for a way forward to expand the Genosolver workflow to additional variant types, such as repeats and CNVs.

## 5. Methods

### Dataset Creation and Data Preprocessing

#### Electronic Health Record Processing

We analysed medical records of 2,108 patients (solved and unresolved cases) at the Center for Human Genetics and Genome Medicine, Uniklinik RWTH Aachen. We extracted 13,566 documents via the local hospital information system and converted them to JSON format before indexing in Elasticsearch. Patient records may include clinical notes, genetic results, demographics, disease descriptions, billing information, or referral letters. Data preprocessing involved noise removal, text excerpting for case summaries, diagnostic result redaction, and German language normalization using methods from the Python standard library before bulk upload to Elasticsearch (1,000 documents per batch). Finally, we filtered out irrelevant phenotype data, leaving 3,456 relevant records classified into structured short descriptions from medical reports or unstructured case summary protocols.

### Curation of genetic dataset

The 233 WES and WGS cases analysed at the Center for Human Genetics and Genomic Medicine at the Uniklinik RWTH Aachen were processed using an internal pipeline^55^, employing KGGSeq^56^ (variants below a 0.75% frequency threshold) and the DRAGEN platform for sequencing analysis. The output comprised annotated Excel files of indels and SNVs from annotation periods in 2023-2025. Variants meeting specific criteria were selected: OMIM phenotype annotations, non-benign ClinVar classifications (pathogenic or likely pathogenic), truncating mutations with frameshift, stop gain effects, splice-donor or splice-acceptor variant, high CADD scores above 20, and homozygous (or compound heterozygous) recessive variants.

### Machine translation

Not all existing methodologies (e.g., PhenoBERT) can handle raw German text for phenotype extraction or conversion into associated HPO terms. Therefore, a multilingual transformer model (ModelSpace/GemmaX2-28-9B-v0.1) (Gemma with 9 billion parameters with 4-bit quantization using the BitsAndBytesConfig library), has been deployed locally, for machine translation. For translating of the full German input text, we have considered a maximum of 2,000 new tokens. For translating the model’s extracted phenotypes (patient clinical features), we have considered a maximum of 512 new tokens. At the end, along with the raw German text, a separate version of the translated descriptions has been stored.

### Summarization of medical notes

From the previously filtered clinical notes described as second category of documents, our primary objective is to extract all the patient’s demographic information and family history from the unstructured text. Recent progress in LLMs shows that zero-shot prompting can improve system performance when the instructions for roles, tasks and reasoning are clear^57^. To summarize the medical documents for a patient, we applied an open-source SLM model (Llama3.1-8B-Instruct^36^) deployed via Hugging Face^58^, using prompt engineering techniques such as Chain-of-thoughts (CoT)^59^ and Tree-of-thoughts (ToT)^60^ to identify four crucial data points summarized in a paragraph. These four crucial data points, packed into a paragraph that we call medical notes, are considered alongside LLM-extracted phenotypic features. Overview of the prompt has been provided in the supplementary document, as supplementary table 1.

### Building a Literature Knowledge Base with Open-Source Datasets

We downloaded all 950 chapters from the GeneReviews gene_NBK1116 database, selecting those pertinent to gene-phenotype associations and genetic conditions. Using PyMuPDF in Python for text extraction, we extracted clinical characteristics and chapter notes content per chapter. This process aimed at facilitating identification of gene-phenotype correlations. We then applied the Named Entity Recognition (NER) model (pruas/BENT-PubMedBERT-NER-Gene) to extract all genes from text within clinical characteristics and notes sections. We extracted gene names from the chapters and compiled them using data from HPOJax’s genes_to_phenotype dataset, filtered against OMIM reference lists. Employing a hybrid prompt with LLM techniques, we determined associations between these genes and phenotypic features within specified texts. Our approach classifies gene-phenotype relationships as rank one for direct correlations or rank two when part of differential diagnoses. The methodology also accounts for contextual nuances such as negation while merging text evidence into key findings, alongside optional fields like age, sex, prevalence, inheritance patterns, mutation types, ethnicity, severity, clinical manifestations, and comorbidities where available. We employed the DeepSeek V3.1 model (deepseek/deepseek-chat-v3.1) via OpenRouter, a significant hybrid reasoning model with 671 billion parameters and 37 billion active ones. The model generates responses incorporating gene names, phenotypic features, priority levels, key findings, age, sex, possibility of occurrence, inheritance patterns, mutation types, ethnicity correlations, severities, clinical manifestations, and associated comorbidities in a structured JSON format based on input from GeneReviews’ 102,899 gene-phenotype rows. A generated example for this can be found in the supplementary material. The prompt involved in the generation of this dataset can be found in Supplementary Table 7.

Another evidence source was the Universal Protein Resource (UniProt), which served as source of literature evidence for establishing gene-disease associations. It integrates and cross-references information from a multitude of extensive and diverse sources, providing the comprehensive collection of protein sequences and functional annotations available^61,62^. It is both thorough and freely accessible, gathering gene-disease association data through manual curation of scientific literature and integration with specialized external databases. These external sources include peer-reviewed literature, OMIM, ClinVar, and repositories such as Orphanet^63^ and genome databases. For the research, we have compiled a total of 12,412 instances of gene involvement in disease mapping, with 12,260 unique genes identified.

### Exomiser

For the 233 cases the Exomiser 14.0.0 version was utilized, in conjunction with the 2406 dataset release, which mirrored the available for the date the original analysis was performed. Exomiser (14.0.0) was run with the standard settings and the WES panel provided by the Exomiser team as well as the expert assigned HPO terms. For the 1,875 cases the Exomiser 15.0.0 version was utilized, in conjunction with the 2512-dataset release, as it has been demonstrated that this combination is especially useful for reanalysis projects. Exomiser (15.0.0) was run with the standard settings and the WES panel provided by the Exomiser team as well as the expert assigned HPO terms.

### Genosolver Phenotype Extraction using LLM

The Phenotype Extraction Module of the Genosolver LLM utilized pre-processed, translated documents. Initially, we loaded local SLMs optimized for GPU operation via Hugging Face’s quantization capabilities. To prevent network calls during instantiation, both tokenizer and model were set with local_files_only = True. The input text was pre-processed by removing leading and trailing whitespace before constructing a chat prompt that merged system instructions (hybrid prompt) with user-provided medical notes. The tokenizer produced a sequence using the chat template, which was subsequently processed to extract only patient clinical information. This data served as phenotypic features for each case in our dataset, with Llama model demonstrating superior performance for this task. Additionally, we employed the PhenoBERT model, a BERT-base encoder designed for HPO term identification. The process involves loading an HPO hierarchy and associated models: fast-text embedding, CNN classifier, Stanza clinical NER (i2b2 corpus), and a specialized BERT classifier to assess the relevance of text segments in relation to specific HPO terms. A two-level CNN generates candidate lists for each note’s phrases extracted via NER. Candidates are ranked using fast-text embeddings and CNN, with subsequent refinement by BERT. Annotations are saved as specified. The supplementary section details the prompt for the LLM-based phenotype extraction (supplementary table 2) and provides details about the small language models and quantisation configurations employed for module phenotype extraction (supplementary table 3).

### Genosolver Rare Disease Diagnosis - Vector Store and MMR

To mitigate the computational burden of ranking all genetic variants, we implemented an intermediate filtering step using patient phenotypic features for coarse classification. We propose a system employing vector storage to generate possible diagnoses based on phenotypic features and facilitate subsequent variant prioritization. The workflow commences with a BERT^64^-based sentence transformer (msmarco-bert-base-dot-v5), generating embeddings of patient phenotypic features, which are then queried against an FAISS^65^ library’s vector store containing curated genotype-phenotype relations for prioritized variant analysis. We utilized the following dataset to construct the vector store: HPO-Jax belongs to the HPO project and provides a collection of standardized phenotypic features, in the form of HPO terms and established Gene-HPO links, which highlight known genes that are associated with certain phenotypic features. We obtained all available HPO terms and their synonyms which were again embedded again by the previously introduced BERT^64^-based sentence transformer. The embeddings of individual terms and their synonyms were aggregated by producing a mean embedding. To prevent the situation that only highly similar phenotypic features are retrieved from the vector storage, for which more terms exist, compared to specific phenotypic features, we utilized the Maximal Marginal Relevance (MMR)^66^ technique to retrieve relevant and diverse phenotypic features from the vector storage. For the retrieved phenotypic features subsequentially their known gene-phenotypic features are obtained. We utilized a compact language model (Mistral-7B-Instruct-v0.1) to classify clinical attributes prior to diagnosing rare diseases using vector store and MMR strategies. The model assessed the provided list for phenotypic features or HPO terms, triggering gene list suggestion upon detection of relevant clinical information.

### Genosolver Variant Prioritization

Finally, the previously prepared input data were combined for the purpose of variant prioritization utilizing different LLMs and LRMs. Using the provided context with a chain-of-thought based hybrid prompt, we generate a report that includes extracted clinical data, key evidence from literature, confidence assessment, comparative analysis, and importantly, ranked genes. Overall, we have compared the following models:

1. deepseek-ai/DeepSeek-R1-Distill-Llama-70B (max_new_tokens=2500, top_k = 10, top_p = 0.7, temperature = 0.0, repetition_penalty=1.1, do_sample=False)
2. OpenMedZoo/MedGo (max_new_tokens=2500, top_k = 10, top_p = 0.7, temperature = 0.0, repetition_penalty=1.1, do_sample=False).
3. unsloth/Llama-3.3-70B-Instruct-bnb-4bit (max_new_tokens=2500, top_k = 10, top_p = 0.7, temperature = 0.0, repetition_penalty=1.1, do_sample=False).
4. GPT OSS 120B using low reasoning (max sequence length = 12000, max new tokens = 2048, temperature = 0.6, top_p = 0.9, do_sample = True)
5. GPT-5.1-Reasoning (Reasoning capabilities: low, medium and high)

Additional information regarding the model architecture of the utilized models can be found in supplementary table 4.

The previously extracted candidate gene list is utilized to enrich the input data for this model with evidence from the literature resources (GeneReviews^67^ and Uniprot^62^). A sentence transformer model (MedEmbed-large-v0.1) generates semantic embeddings from the text, which are then processed using a Maximal Marginal Relevance algorithm to create an overview of genetic evidence based on GeneReviews data for specific genes and UniProt database information related to candidate gene lists in patients’ profiles. This comprehensive context (which was provided in different combinations, depending on the experiment) includes phenotypic features, family history, patient medical histories, HPO terms, and the extracted candidate gene list. A summary of the experimental set ups can be found in supplementary table 5. The prompt utilized in these experiments can be found in supplementary table 6.

### Reanalysis

The analysis employed Exomiser version 15.0.0 and the 2512 dataset to extract unstandardized phenotypic features from available medical documents, as previously described. Using standard settings in Exomiser with WES presets and HPO terms, expert-assigned HPO terms were applied when possible; otherwise, derived HPO terms based on identified phenotypes. From the ranked variants by Exomiser, we retained: Known Pathogenic or likely pathogenic (Exomiser^41^ and ClinVar^46^) variants with OMIM^68^ phenotype associations. Additionally, VUS variants with a variant score above 0.7 for variants with new gene-phenotype links in OMIM^68^, compared to the previous manual analysis. These were evaluated using Llama within Genosolver workflow. A total of 1,875 cases underwent additional review to understand previously unreported variant occurrences.

### Computational Resources Used in This Study

For on premise analysis two GPU systems were used. One system was equipped with a 24GB VRAM RTX 3090, which was utilized to perform the computational lighter tasks, such as translation. Additionally, a Nvidia H100 DGX system equipped with 8 H100 80GB VRAM GPUs was leveraged. Unless stated otherwise, all model evaluations were conducted locally to maintain data privacy and minimize latency. The experiments utilizing the GPT-5.1 models were conducted using Azure AI Foundry with GDPR-compliant EU data residency.

## Supporting information

Supplementary Chapters

## Data Availability

The code developed for this study will be made publicly available on GitHub upon publication.

## 6. Abbreviations

CNVs: Copy Number Variants
LLMs: Large Language Models
LRMs: Large Reasoning Models
HPO: Human Phenotype Ontology
MMR: Maximal Marginal Relevance
NER: Named Entity Recognition
NGS: Next-Generation Sequencing
GQA: Grouped-Query Attention
RoPE: Rotary Position Embeddings
SLM: Small Language Model
SVs: Structural Variants
SWA: Sliding Window Attention
YaRN: Yet another RoPE extensioN
MoE: Mixture of Experts
VUS: Variants of Uncertain Clinical Significance
WES: Whole Exome Sequencing
WGS: Whole Genome Sequencing
CoT: Chain-of-Thought Prompting
ToT: Tree-of-Thought Prompting

## 7. Declarations

### Ethics approval and consent to participate

This study was approved by the ethics committee at the Medical Faculty of RWTH Aachen University (Approval No: EK 302/16 & EK 126/22). Informed consent was obtained from all participants prior to their inclusion into the study. Patient-Level data was de-identified.

### Consent for publication

All authors approved the final version of the manuscript for publication.

### Availability of data and materials

The code developed for this study will be made publicly available on GitHub upon publication.

## Acknowledgements

**None**

## Authors Contribution

**TI:** Conceptualization, Methodology, Software, Writing - Original Draft, Visualization, Formal analysis, Investigation **MD:** Software, Methodology, Formal analysis, Investigation **ZZ:** Conceptualization, Methodology, Software, Formal analysis, Investigation **MB:** Conceptualization, Methodology, Software, Validation, Resources, Data Curation, Formal analysis, Investigation **DB:** Conceptualization, Methodology, Resources, Data Curation **AL:** Resources, Data Curation **EL:** Resources, Data Curation **LM:** Resources, Data Curation **JS:** Resources, Data Curation **PW:** Resources, Data Curation **NG:** Resources, Data Curation **ES:** Resources, Data Curation **RK:** Resources, Data Curation **SD:** Resources, Data Curation **LF:** Resources, Data Curation **JR:** Software Resources, Data Curation **Robin M:** Software Resources, Data Curation **EP:** Project administration **HZ:** Resources, Data Curation **NH:** Resources, Data Curation **KE:** Resources, Data Curation **TE:** Resources, Data Curation **RM:** Conceptualization Resources, Data Curation **FK:** Conceptualization, Methodology, Software **ME:** Conceptualization, Methodology, Validation, Resources, Data Curation, Supervision, Project administration **IK:** Conceptualization, Methodology, Validation, Resources, Data Curation, Supervision, Project administration **JK:** Conceptualization, Methodology, Software, Validation Resources, Data Curation, Writing - Original Draft, Visualization, Supervision, Project administration, Funding acquisition, Formal analysis, Investigation **All Authors:** Writing - Review & Editing

