## Supplementary Chapters for "Genosolver: Rare Disease Diagnosis through Holistic Integration of Unstructured Clinical Narratives Using Large Language and Reasoning Models"

### Supplementary Tables:

Supplementary Table 1: Description of the prompt utilized in the summarization task.

| Section | Functions |
| --- | --- |
| Role | It sets the model’s persona as a clinical text analyst. |
| Task | It specifies the summarization objective and limits the output to patient history, family history, age, and sex. |
| Input | Inserts the raw clinical text for processing. |
| Reasoning Steps - CoT^1^ | This step identifies all sentences and phrases describing the patient’s history, family history, age, and sex. It detects and excludes sections related to examination findings, test results and diagnostics. Finally, it reformulates the extracted information. |
| Evaluation Steps – ToT^2^ | In this step, it verifies that no diagnostic or test details remains; so, removes any that are found in the text. Review that each key factor is included if present in the text. Finally, it evaluates the clarity and focuses on the final summary. |
| Output format | A concise summary containing only four key elements. |

Supplementary Table 2: Overview of the prompt utilized in the LLM-based phenotype extraction task.

Prompt

You are a medical information extraction assistant. Your role is to extract relevant clinical information from free-text patient case descriptions.

The input text may include:

    - Phenotypic details (signs and symptoms)

    - Differential diagnoses

    - Associated genes and genetic variants

    - Information about both the patient and their relatives

Your task is to:

    - Extract and organize medically relevant details **only for the patient**

    - Identify which differential diagnosis is most consistent with the information given

    - Extract relevant genetic findings for the patient

    - Clearly explain why other differentials are less likely

    - Do not make any medical assumptions — only extract, match, and organize what's explicitly stated in the text

IMPORTANT:

    - If the text says the patient is a **carrier** of a variant, do **not assume** they have the disease associated with it

    - Only include gene-disease associations when there is **clear evidence of clinical relevance in the patient**

    - Exclude variants or findings that apply **only to relatives** unless the patient also shows relevant symptoms

    ---

    1. Most likely diagnosis (as explicitly supported by the text): <letter>. <full diagnosis name>

    2. Evidence from the text:

    - Patient clinical features:

        - <feature 1>

        - <feature 2>

        - <...>

    - Patient gene variants (exclude carriers unless clinically relevant):

        - <gene 1>

        - <gene 2>

        - <...>

    - Gene-disease associations (from text, only if relevant to the patient):

        - <gene> is associated with <disease>

        - <gene> is associated with <disease>

    - Differential diagnoses mentioned:

        - A. <name>

        - B. <name>

        - ...

    3. Matched disease-gene pair (most supported): <disease> – <gene>

    4. Confidence score (based only on strength of match in text): <score from 0–1>

    ---

Table 3: Description of SLM models used in the module Genosolver LLM-based phenotype extraction.

| Model | Variant | Parameters | Approx.  Size | Precision | Architecture | Context Length |
| --- | --- | --- | --- | --- | --- | --- |
| PhenoBERT^3,4^ | default | ~110 M | ~440 MB | FP32 | Two-level CNN + BERT-Base (Encoder), bidirectional self-attention | 512 |
| Llama 3.1 8B Instruct^5^ | Full | 8.0 B | ~16 GB | BF16 | Decoder-only Transformer, GQA, RoPE | 128K |
|  | Quantised | 8.0 B | ~8 GB | NF8 |  |  |
| Gemma 7B^6^ | Full | 8.5 B | ~17 GB | BF16 | Decoder-only Transformer, Multi-Head Attention, RoPE | 8,192 |
|  | Quantised | 8.5 B | ~8.5 GB | NF8 |  |  |
| Mistral 7B Instruct v0.1^7^ | Full | 7.3 B | ~14.5 GB | BF16 | Decoder-only Transformer, GQA, SWA, RoPE | 8,192 |
|  | Quantised | 7.3 B | ~7.3 GB | NF8 |  |  |

Supplementary Table 4: Summary of the LLMs and LRMs used for the module variant prioritization

| Model | Lineage | Total  Parameters | Active  Parameters | Precision / Quantisation | Architecture | Context Length |
| --- | --- | --- | --- | --- | --- | --- |
| DeepSeek-R1-Distill-Llama-70B, Distilled^8^ | Llama 3.3-70B-Instruct, fine-tuned on DeepSeek-R1 reasoning data | 70B | 70B | BF16 | Decoder-only Transformer, GQA, RoPE | 128K |
| MedGo | Qwen3^9^-32B, two-stage medical SFT on multi-source clinical corpora | 32.8 B | 32.8 B | BF16 | Decoder-only Transformer, GQA, RoPE | 32K |
| Llama 3.3 70B Instruct, 4bit, Reasoning (low)^5^ | meta-llama/Llama-3.3-70B-Instruct, pre-quantised by Unsloth | 70 B | 70 B | NF4 | Decoder-only Transformer, GQA, RoPE | 128K |
| GPT OSS 120B^10^ | Trained from scratch | 117B | ~5.1 GB | MXFP4 | Decoder-only Transformer, MoE, GQA, SWA, RoPE, YaRN | 128K |

Supplementary Table 5: Experiments performed for variant prioritization

| Experiments | Input structure |
| --- | --- |
| Experiment 1 | - The unstandardized phenotypic features are provided as input. - The relevant gene list is derived by performing a vector search utilizing the unstandardized phenotypic features. - Summarized medical notes containing age, sex, family history are provided as an additional input. - Context from the literature evidence database (GeneReviews and UniProt). |
| Experiment 2 | - The expert HPO terms from the subject-matter experts. - The relevant gene list is derived by performing a vector search utilizing the expert HPO terms. - Context from the literature evidence database (GeneReviews and UniProt). |
| Experiment 3 | - The phenotype derived HPO terms are utilized as an input.   a. Phenotypic features extracted using the proposed method.   b. Applied PhenoBERT approach to the phenotypic features to received HPO ids.   c. Converted HPO ids to HPO terms using the Ontology Lookup Service (OLS) API. - The relevant gene list is derived by performing a vector search utilizing the derived HPO terms. - Context from the literature evidence database (GeneReviews and UniProt). |
| Experiment 4 | - The unstandardized phenotypic features are provided as input. - Integrating Exomiser’s ACMG classification:   a. The relevant gene list is derived by performing a vector search utilizing the unstandardized phenotypic features   b. Genes are discarded if the corresponding variants are classified benign or likely benign by Exomiser. - Summarized medical notes containing demography information. - Context from the literature evidence database (GeneReviews and UniProt) |
| Experiment 5 | - The unstandardized phenotypic features and the expert derived HPO-terms are provided as input - Integrating Exomiser’s ACMG classification:   a. The relevant gene list is derived by performing a vector search utilizing the unstandardized phenotypic features   b. Genes are discarded if the corresponding variants are classified as benign or likely benign by Exomiser   c. Filter gene list further based on Exomiser’s gene variant score above 0.7. - Summarized medical notes containing age, sex, and family history. - Context from the literature evidence database (GeneReviews^11^ and UniProt^12^) |

Supplementary Table 6: Description of the prompt utilized in the variant prioritization task.

Prompt:

You are a human genetics expert, PhD/MD level, tasked with identifying the most disease-causing genetic variant.

Patient Data (if available):

- Selected Phenotypes (English): {phenotypes}

- Relevant Genes: {genes}

- Medical Notes: {medical_notes}

Literature Evidence:

{context_str}

Task Instructions:

1. Extract Clinical Information:

- Determine patient's age, sex, and other relevant clinical information from the medical notes or context.

- List all extracted info clearly.

2. Key Evidence Extraction (maximum 300 words):

- for each Gene, identify and summarize the most relevant facts based on given *Selected Phenotypes* and *Relevant Genes* from the literature evidence.

- Each gene, single summarized line

3. Comparative Reasoning:

- Rank *all Genes* (e.g., 1st, 2nd, 3rd, ......)

- Explain why one gene/variant is more likely than others, considering phenotypes, age, sex, and clinical info.

4. Final Decision:

- Identify the single most disease-causing variant.

- Justify your choice using evidence and clinical reasoning.

5. Structured Summary:

- Provide Ranked list for *all* genes. *STOP* after 10 genes.

- Final Most Disease-Causing Variant

- Comparative Reasoning Summary

Guidelines:

- Only use the provided phenotypes, genes, medical notes, and literature evidence.

- Provide step-by-step reasoning and make your justification explicit.

- Use bullet points or tables for clarity.

- Provide concise, evidence-based explanations.

- Avoid producing JSON format in the response.

Question: {question}

Answer:

Supplementary Table 7: The prompt for gene-phenotype association extraction, applied to the GeneReviews chapters dataset.

Prompt:

You are a professional biomedical researcher specializing in gene–phenotype associations.

Extract relationships between the gene **{gene_name}** and phenotypes

(diseases, clinical traits, observable characteristics) **only from the text below**.

Instructions:

1. Identify all phenotypes mentioned in the text with the gene **{gene_name}** and normalize their names.

2. Assign a priority for each phenotype mentioned together with the gene **{gene_name}**:

- Rank 1: phenotype related to the gene **{gene_name}** should be labeled Rank 1

- Rank 2: phenotype related to different gene names differential diagnosis should be labeled Rank 2

3. Consider context and negation. Ignore explicitly negative or uncertain associations.

5. Summarize key evidence from the text in the "Key findings" field.

6. Extract optional fields if available:

- age, sex, possibility/prevalence, inheritance_pattern, mutation_type, ethnicity, severity, clinical_manifestation, comorbidities, treatment.

- If a field is missing, leave it empty.

7. **Output format:** Use structured free text, **one phenotype per block**, like this:

---

Gene: {gene_name}

Phenotype: ...

Priority: ...

Key findings: ...

Age: ...

Sex: ...

Possibility: ...

Inheritance pattern: ...

Mutation type: ...

Ethnicity: ...

Severity: ...

Clinical manifestation: ...

Comorbidities: ...

Treatment: ...

---

8. Process long texts in chunks if needed. Do not truncate your output. If all phenotypes cannot fit in a single response, output them sequentially in the same structured format without skipping any.

**Important:**

- Only report phenotypes explicitly mentioned in the text. Do not invent new ones.

- Keep the output strictly in the structured format above, with no extra commentary or JSON wrapping.

Text for extraction:

"{text_content}"

Supplementary Table 8: Description of the prompt utilized in the variant prioritization for the CNV task.

Prompt:

You are a human genetics expert, PhD/MD level, tasked with identifying relevant genetic variants for specific phenotypes.

Patient Data (if available):

- Selected Phenotypes (English): {phenotypes}

- Relevant Genes affected by a copy number variation: {genes}

- Medical Notes: {medical_notes}

Literature Evidence:

{context_str}

Task Instructions:

1. Extract Clinical Information:

- Determine patient's age, sex, and other relevant clinical information from the medical notes or context.

- List all extracted info clearly.

2. Key Evidence Extraction (maximum 300 words):

- for each Gene, identify and summarize the most relevant facts based on given *Selected Phenotypes* and *Relevant Genes* from the literature evidence.

- Each gene, single summarized line

3. Comparative Reasoning:

- Rank *relevant Genes* (e.g., 1st, 2nd, 3rd,......)

- Explain why one gene/variant is relevant or not relevant to the phenotype, considering phenotypes, age, sex and clinical info.

4. Final Decision:

- Identify multiple relevant genetic variants.

- Justify your choice using evidence and clinical reasoning.

5. Structured Summary:

- Provide Summary wether genes affected by the copy number variant can explain the patient’s phenotype

- Identify multiple relevant genetic variants.

- Comparative Reasoning Summary

Guidelines:

- Only use the provided phenotypes, genes, medical notes, and literature evidence.

- Provide step-by-step reasoning and make your justification explicit.

- Use bullet points or tables for clarity.

- Provide concise, evidence-based explanations.

- Avoid producing JSON format in the response.

Question: {question}

Answer:

Supplementary Figures:

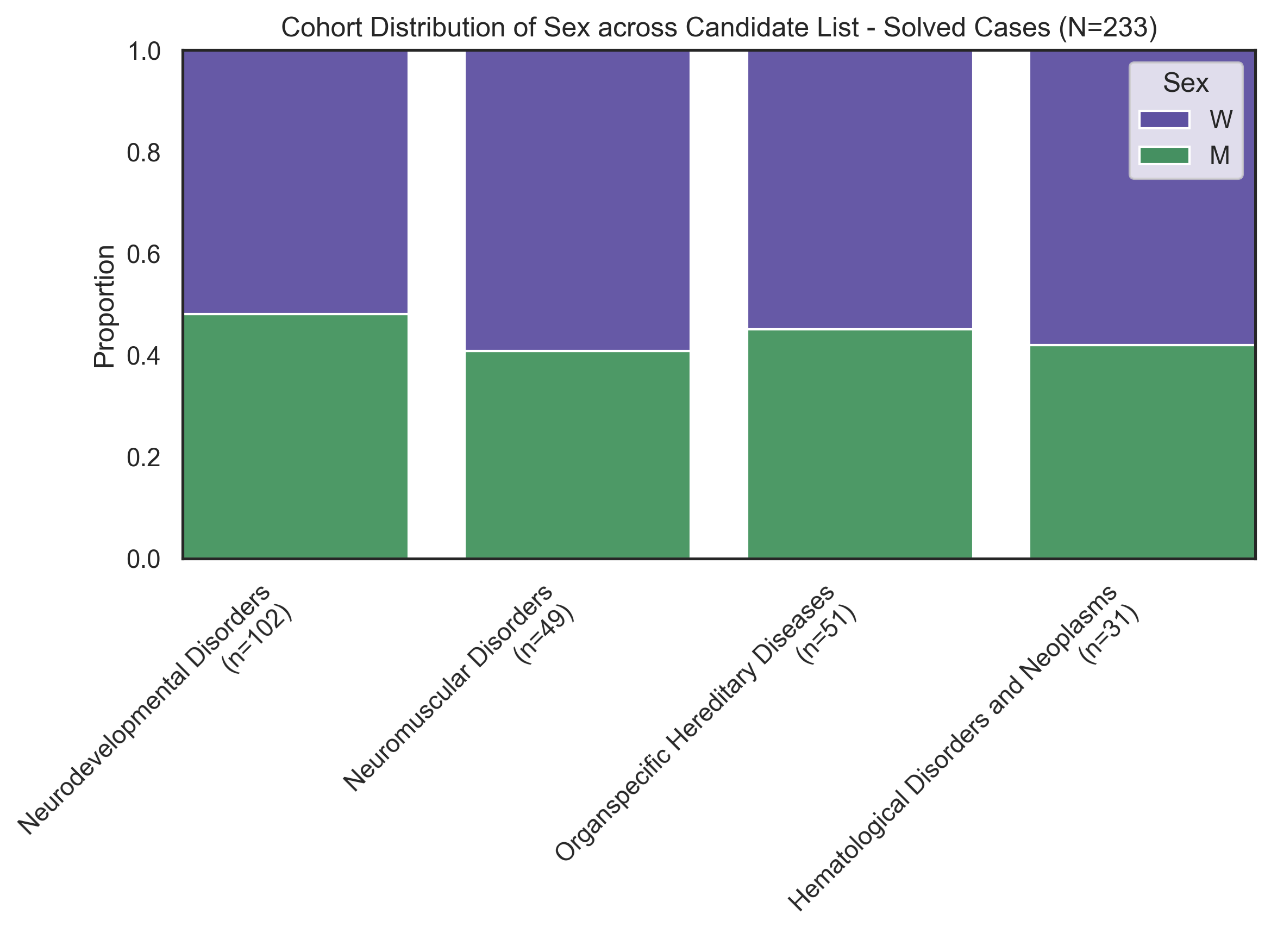

*Supplementary Figure 1: Distribution of the sex and grouped rare disease categories for the 233 molecularly solved patients with rare diseases.*

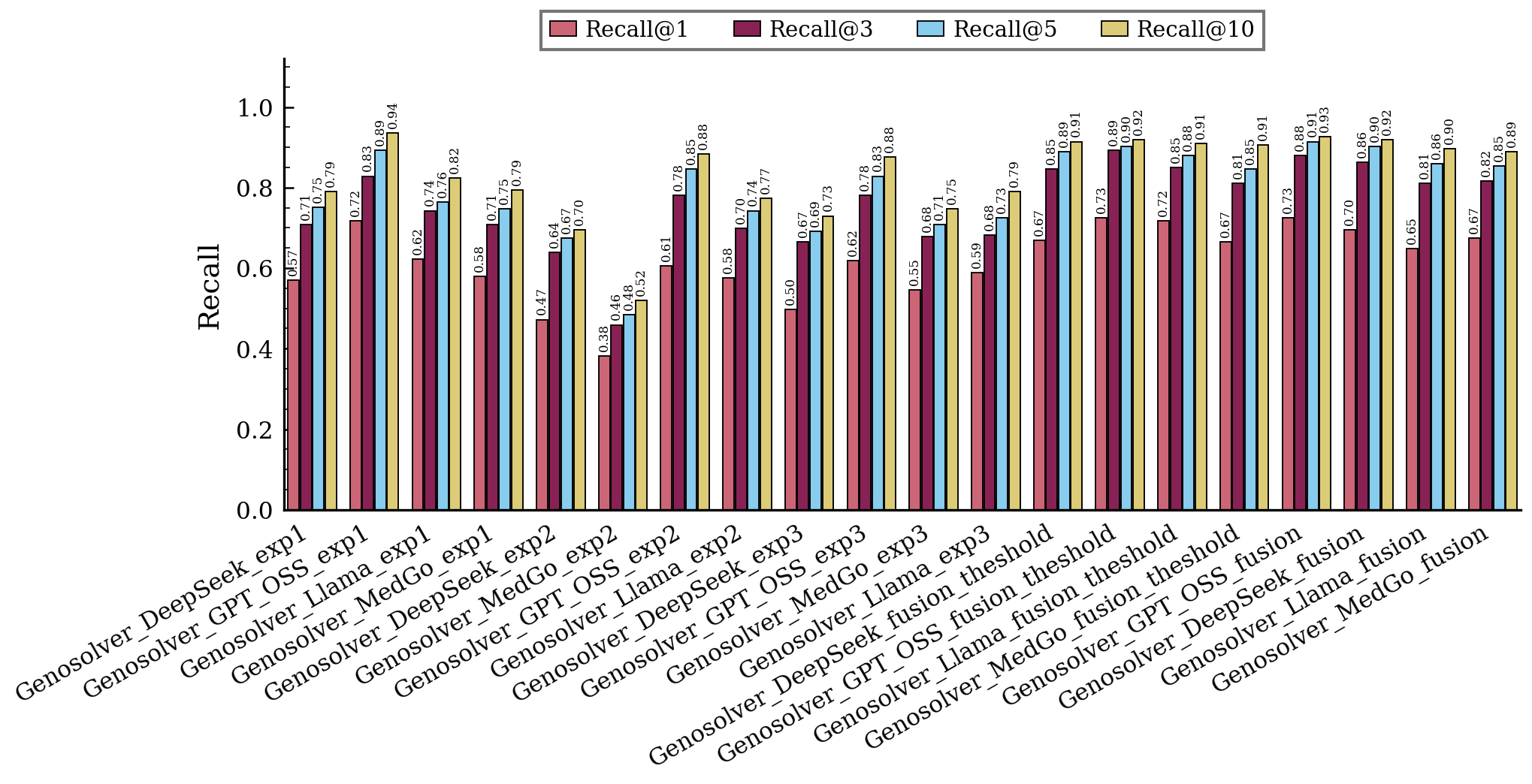

*Supplementary Figure 2: Summary of Recall across different LLMs for Experiments 1–5 is provided. Each bar represents recall in one experiment corresponding to values of 1, 3, 5, and 10 accordingly.*

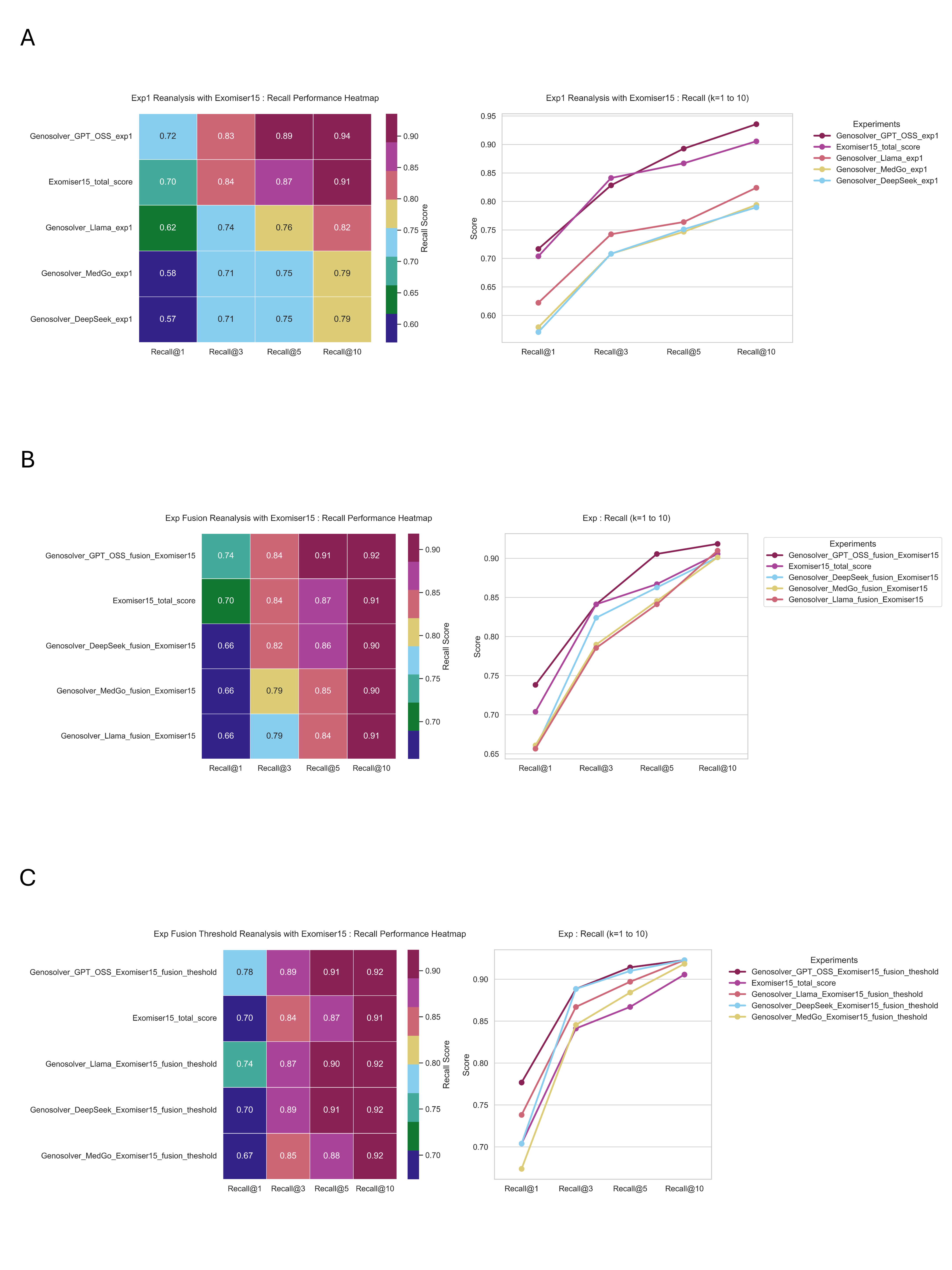

*Supplementary Figure 3: Head-to-head comparison between Genosolver and Exomiser 15.0.0 for the best performing model. A) In Experiment 1, phenotypic features have been extracted using Genosolver LLMs, then differential diagnosis has been performed, and later Genosolver variant prioritization has been applied. In the figure, performance of the Genosolver approaches has been compared with the total score of Exomiser 15.0.0 experiment. B) Phenotypic features* *Extracted Using Genosolver LLMs, Differential Diagnosis Performed, Candidate Genes Filtered via Exomiser 15.0.0 ACMG Classification, and Genosolver Variant Prioritization Applied, with Performance Compared to Exomiser 15.0.0 total score Experiment. C) Phenotypic features Extracted Using Genosolver LLMs, HPO terms suggested by subject-matter experts, Differential Diagnosis Performed, Candidate Genes Filtered via Exomiser 15.0.0 ACMG Classification and Gene variant scores kept above 0.7 threshold, and Genosolver Variant Prioritization Applied, with Performance Compared to Exomiser 15.0.0 total score Experiment.*

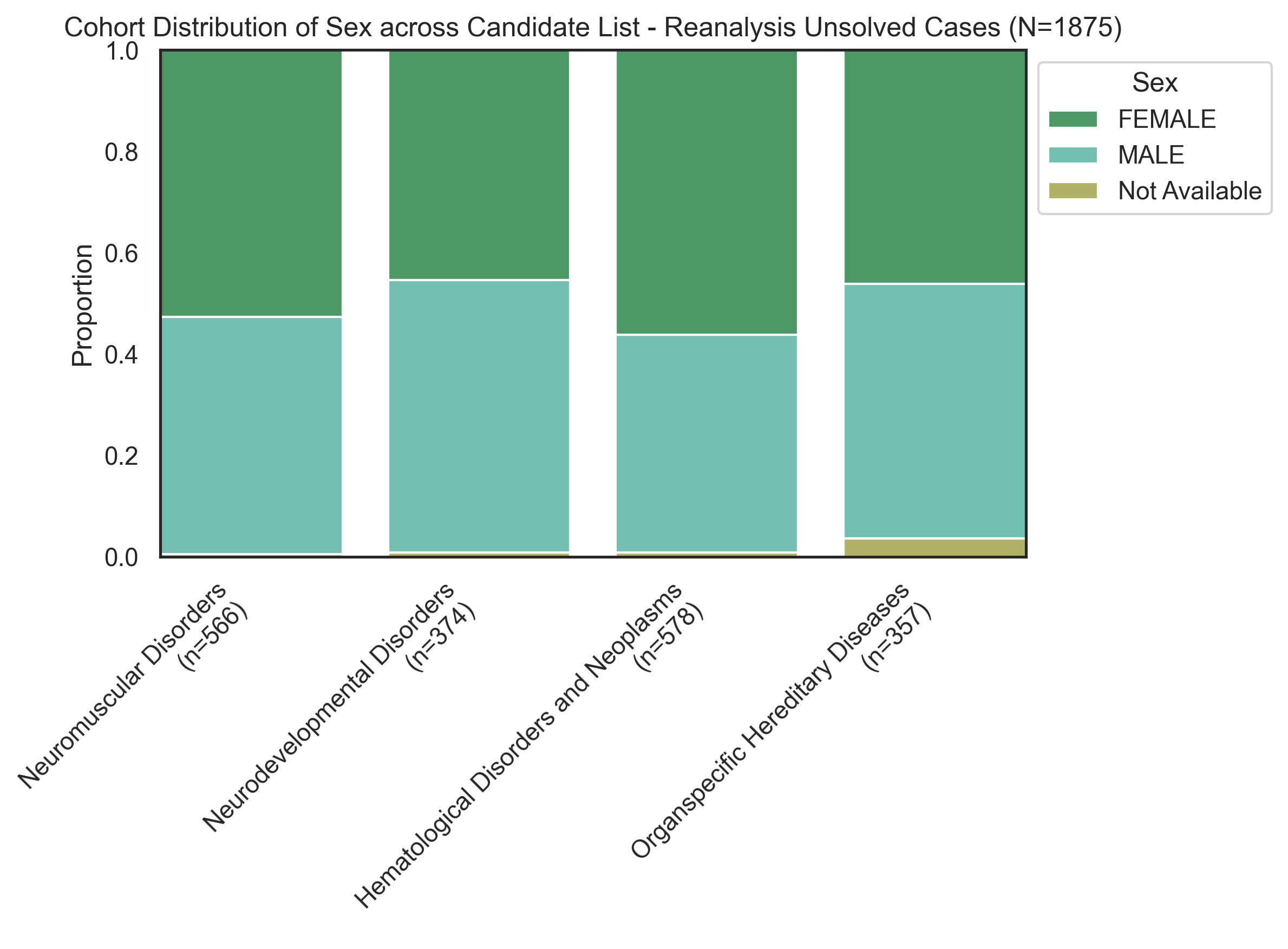

*Supplementary Figure 4: Illustration of the cohort distribution of sex across candidate list for the reanalysis of the unsolved cases (N=1,875) of the years 2023 and 2024 selected for reanalysis.*

### Supplementary Chapter (Phenotype Extractor):

To evaluate the capabilities of several general language models for phenotype extraction , we tested the ability to extract HPO terms from structured and unstructured clinical notes. The performance was compared against the established model PhenoBERT^3^. We evaluated small language models (SLMs) including Llama3.1-8B-Instruct^5^, Gemma-7B^6^ and Mistral7B v0.1^7^, both individually and in combination with PhenoBERT^3^. Nine workflows were evaluated using multiple publicly available datasets (ID-68^13^, the GeneReviews^11^ and the OMIM^14^ dataset from the PhenoBERT^3^ paper). The evaluation metrics included precision, recall, and F1 score, with both micro-averaged (overall performance across all classes) and macro-averaged (average performance per class) values reported. PhenoBERT^3^ demonstrated the best mean performance across the three datasets when used alone, but combining PhenoBERT^3^ with additional SLMs yielded a synergistic effect and further improved performance (Supplementary Figure 3). For example, on the ID-68 dataset, PhenoBERT^3^ achieved F1 scores of 0.85 (micro) and 0.84 (macro), while the best SLM (quantized Llama) achieved 0.81 (micro) and 0.82 (macro). The combination of PhenoBERT^3^ with quantized Llama achieved the highest F1 scores of 0.86 (micro and macro). Similar trends were observed for the OMIM and GeneReviews datasets. Based on these results, we selected the fusion of PhenoBERT^3^ and Llama for downstream analyses.

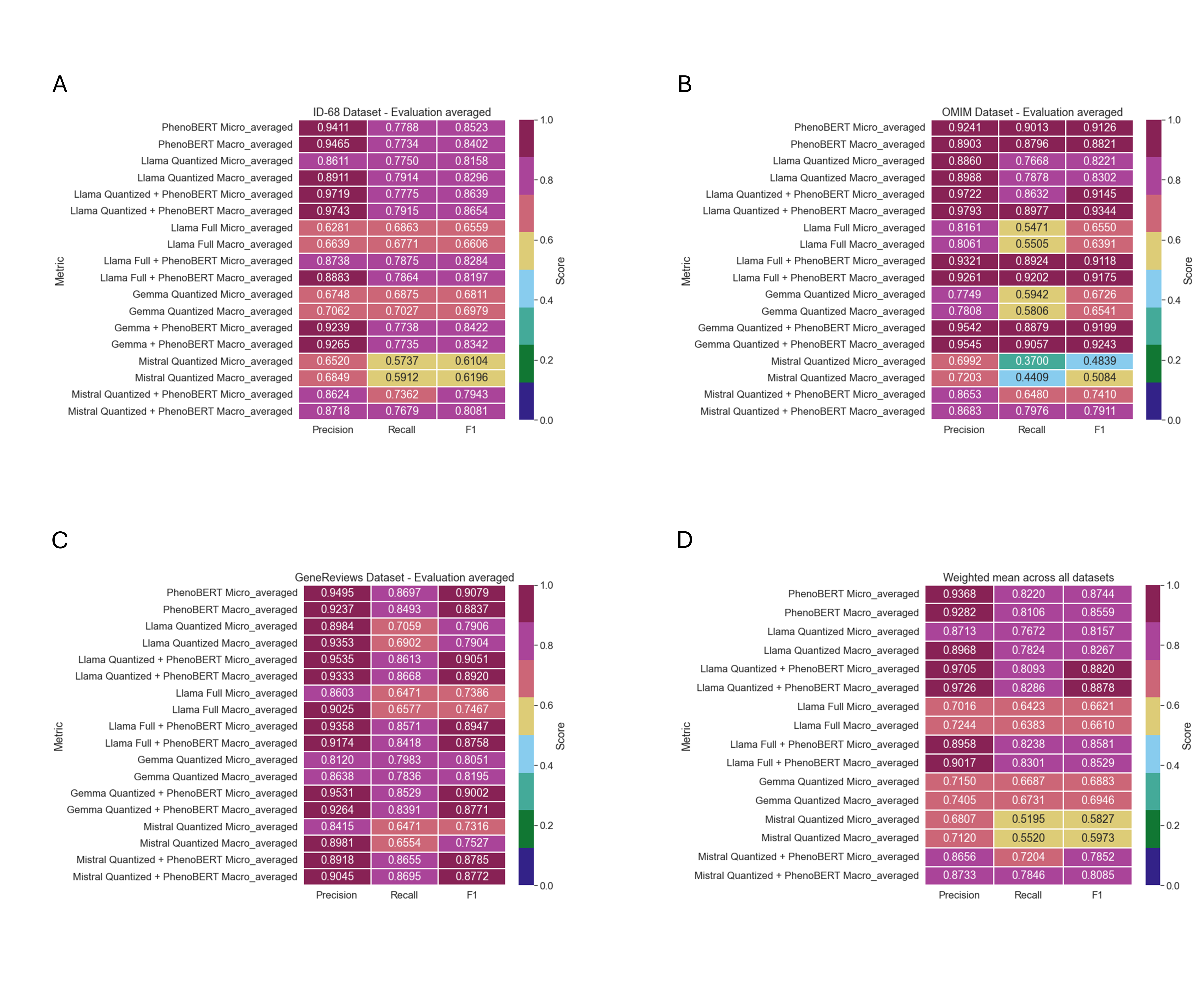

*Supplementary Figure 5: Comparison of the different phenotype and HPO-term extraction strategies. A) Performance evaluation of the PhenoBERT, Llama, Gemma, Mistral for HPO terms recognition on the ID-68 clinical description. Heatmap representing the correlation between predicted phenotypic features* *and expert-annotated ground truth. B) Correlation heatmap comparing the performance of PhenoBERT, Llama, Gemma, and Mistral on HPO term recognition in OMIM clinical research articles (model predictions vs. expert-annotated ground truth). C) Performance evaluation of PhenoBERT, Llama, Gemma, and Mistral for HPO term recognition on GeneReviews clinical descriptions): correlation heatmap of model-predicted phenotypic features vs. expert-annotated ground truth. D) Heatmap illustrating the weighted mean performance scores (Precision, Recall, and F1) across PhenoBERT, Llama, Gemma and Mistral small language models and evaluation metrics, computed proportionally to dataset sample sizes.*

### Supplementary Chapter (Disease Designator):

The second module of Genosolver proposes potential diagnoses based on patient phenotypic features. It functions as a preliminary filter by comparing input genetic information with stored gene-phenotype associations in its vector database to identify relevant matches for further analysis. This process accommodates both structured and unstructured data inputs, facilitating the identification of rare diseases associated with specific phenotypes. As proof-of-concept we used the system to rank potential diagnoses for 233 in-house patients who underwent whole genome or whole exome sequencing and for whom a likely pathogenic (ACMG^15^ class 4) or pathogenic (ACMG^15^ class 5) variant had previously been identified. These cases were subsequently used to evaluate Genosolver's variant prioritization capabilities in the third module. For solved cases, we calculated the reduction factor of this strategy by providing the system with a list of genes containing rare variants (as determined by the in-house pipeline) and the patient's phenotypic features, then receiving filtered gene lists as output. Evaluation criteria included the hit rate (the proportion of cases in which the ground truth gene remained after filtering) and the overall reduction of the gene list. We evaluated three different setups: (1) ranking using unstandardized phenotypic features extracted from clinical notes, (2) ranking using HPO terms assigned by medical experts, and (3) ranking using HPO terms generated by the PhenoBERT-SLM approach based on extracted phenotypic features. The best performance was achieved with the first setup (hit rate 94.3%), followed by expert-assigned HPO terms (90.4%), and finally PhenoBERT-generated HPO terms (89.2%).

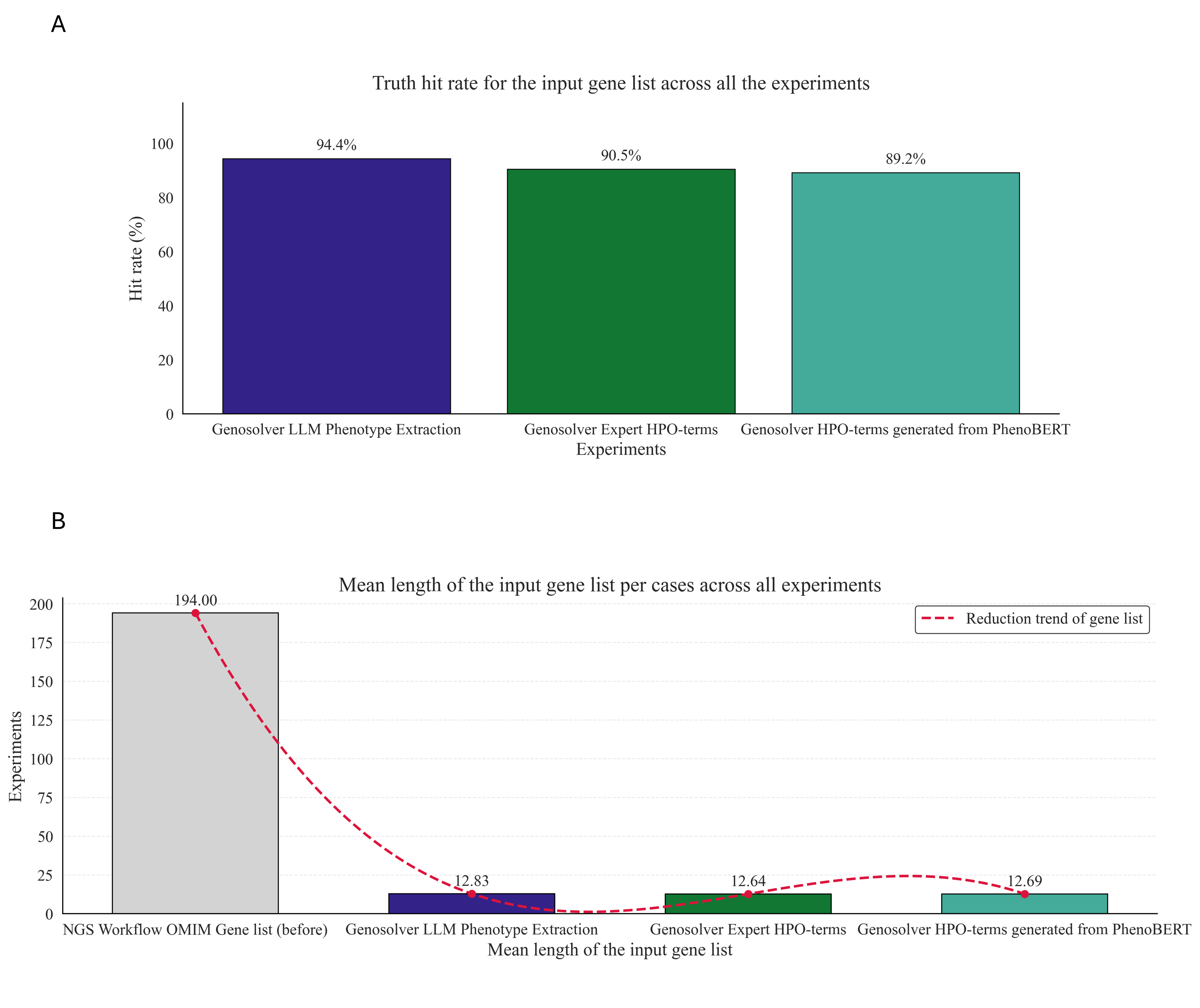

*Supplementary Figure 6: A) The figure illustrates the truth-hit frequency per experiment of the rare disease diagnosis. Here bar indicates the number of cases the ground truth gene was present in the gene list and calculates the hit rate (%). B) Mean length of the input gene list per cases across experiments such as Genosolver LLM phenotype extraction, expert HPO-terms, HPO-terms generated from PhenoBERT (before vs after).*

Supplementary Chapter (Comparison to closed weight models):

An interesting observation we made across the different variant prioritization experiments was that the largest model out of the four utilized models (GPT OSS 120B) was the overall best performing model. While we previously highlighted the possibility of Genosolver to be run in a local set up, we additionally wanted to explore the benefit of larger models. Out of the study cohort we utilized we had the opportunity to use a subset of the cases (n=76) in GDPR-compliant cloud computing environments. Subsequentially, we evaluated this subset utilizing GPT 5.1 with different reasoning settings. On this subset, the best standalone performance was achieved with GPT 5.1 with low reasoning settings, which reached hit rates of 0.71 (k=1), 0.84 (k=3), 0.88 (k=5) and 0.96 (k=10), outperforming the overall best performing GPT OSS model on all hit rates, except the k=10 hit rate. Together this hints to an additional benefit of utilizing even more advanced models.

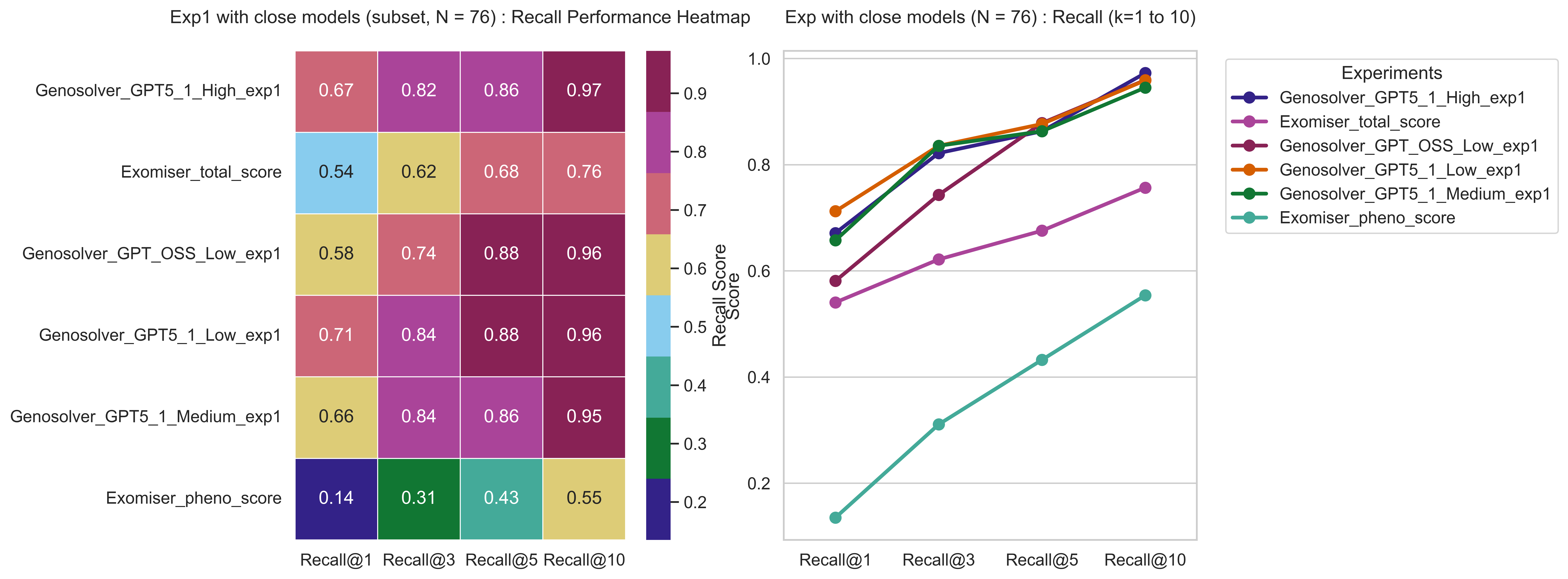

*Supplementary Figure 7: The figure shows the Experiment 1 performance across the close models (subset, N = 76) and compare the results with Exomiser 14.0.0 and best open-weight model for the Experiment1. Here, phenotypic features have been extracted using Genosolver LLMs, then differential diagnosis has been performed, and later Genosolver variant prioritization has been applied.*

### Supplementary Chapter (Performing a confidence analysis for Genosolver):

Here we analysed the rhetorical structure of the best-performing model of Experiment 1 (GPT OSS 120B) with the Genosolver framework. Each final LLM-generated output is gone through text normalisation and segmentation. Such as: It normalises line endings and splits the input into paragraphs. Each paragraph is rolled up into a single string and then tokenized into sentences. Each final LLM-generated output is split into paragraphs and sentences. Every sentence gets two annotations. The first is a rhetorical role such as general, evidence, causal, contrast, hedging, or conclusion determined by matching against a keyword list specific to each role. The function iterates over a priority list of (role, [keyword]) pairs, returns the first matching role. It falls back to “general” if no keyword matches. The second is a confidence score, calculated by words that indicate confidence in the results (e.g., “Most disease-causing variant”, “Evidence”) and words that indicate hedging (e.g., “could potentially”, “Not found”). We also computed a lexical hedging score from uncertainty-related terms and extracted sentence token count and type-token ratio as lexical diversity measure. Token count is obtained by whitespace tokenization, and type-token ratio is calculated as distinct tokens divided by total tokens. In addition, sentence vectorization was performed using scikit-learn^16^ TfidfVectorizer to construct tf-idf matrix across the entire sentences. Moreover, the token count, type-token ratio, and tf-idf vectors are each z-standardised for visual comparison. In summary, this analysis has been developed to review the surface-level understanding of the style of the generated output by the Genosolver variant prioritization stage of the LRM models.

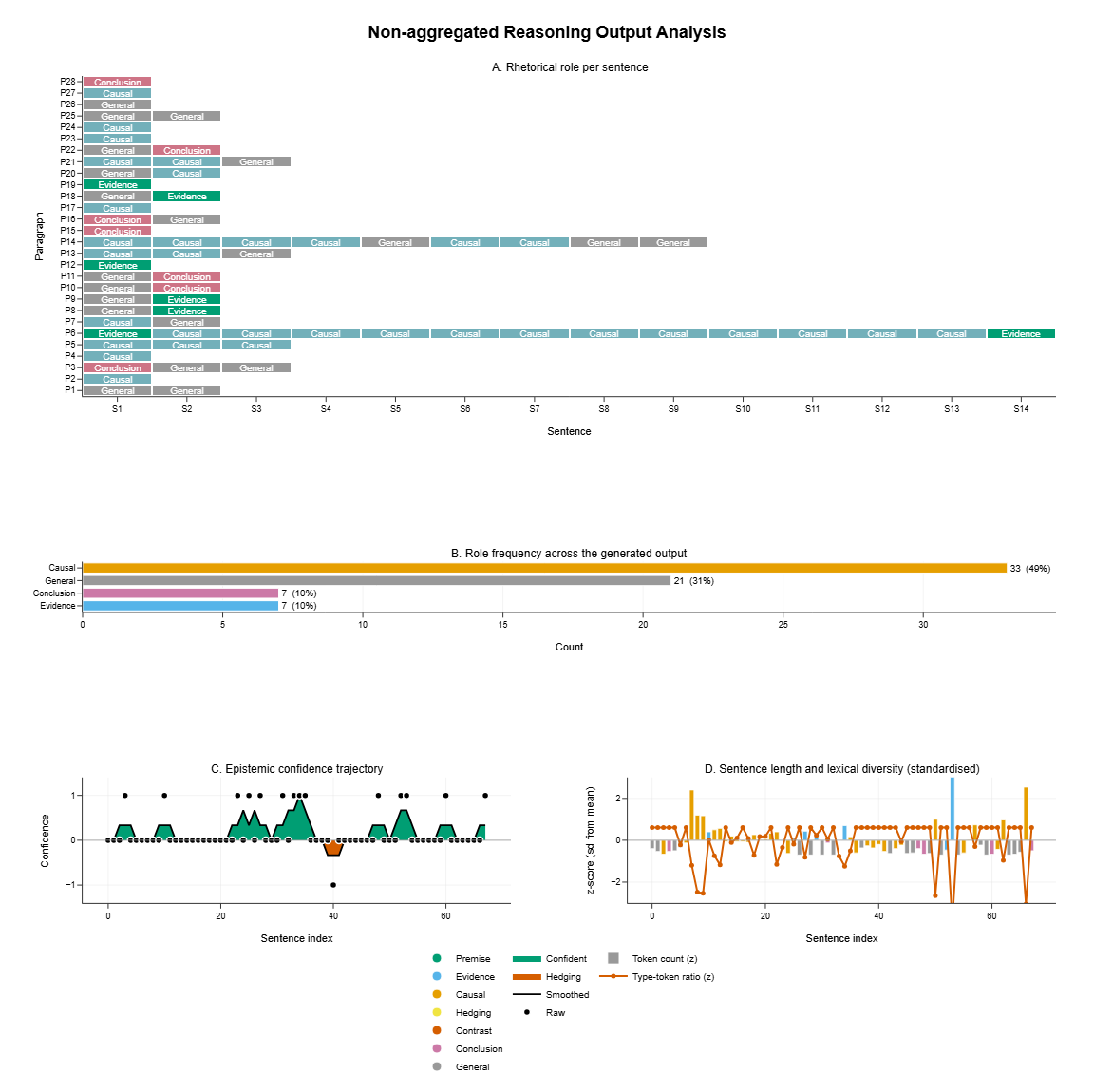

*Supplementary Figure 8: The figure shows the sentence-level discourse and lexical profile of the Genosolver variant prioritization report of a patient, providing clarity on its reasoning features. A) Each sentence’s rhetorical role is indexed by paragraph (rows) and sentence (columns). B) Total counts and percentages for each role are displayed, calculated across all sentences of the specific case. C) Epistemic confidence, ranging from -1 to +1, is shown per sentence. A three-sentence rolling mean is plotted as a black line, with confident and hedging bands also indicated. D) Sentence length and lexical diversity are presented.*

Supplementary figure 6 shows the non-aggregated generated output analysis on surface-level for a particular case of the Genosolver variant prioritization report, where panel (A) illustrates a heatmap with rows showing paragraphs and columns showing sentence position within those paragraphs. It revealed whether the LLM-generated output provides a reasoning structure with causal, general information, evidence, and concluding thoughts. Panel 8B provides the bar graph that quantifies the number of sentences assigned to each role. In this example, the generated output for variant prioritization contained 28 paragraphs. Of these, 31% of the sentences were classified as having a general role, another 49% as having a causal role, 10% as conclusive, and 10% as evidence. Panel 8C visualizes confidence across the generated output, with sentence index on the x-axis and confidence score on the y-axis. A total of seventeen phrases indicating certainty were observed, involving examples like “evidence,” “final,” “most disease-causing variant,” and “clearly,” (e.g., “Final Decision Most disease‑causing variant: *UBA1* (somatic mutation causing VEXAS syndrome)”) besides one instance of hedging language (e.g., “Also perichondritis and vasculitis could be associated with auto-inflammatory diseases like DADA2 (*ADA2* gene not listed)”. Finally, panel D provides a lexical view: for each sentence, it shows the token count and the type-token ratio. Both metrics have been normalized for a shared axis. The figure indicates that the evidence role corresponds to the tallest bar at sentence index 53, signifying that 204 tokens were utilized to characterize the literature evidence derived from GeneReviews and UniProt.

Furthermore, we have analysed all 233 solved cases using the Genosolver Experiment 1 approach with GPT OSS 120B, our best-performing model, incorporating its variant-prioritization results. The aggregated output revealed a total of 6471 paragraphs and 18062 sentences. The overall confidence phrase was 3180, with 14715 neutral phrases and only 167 exceedingly small number of hedging phases identified. This disparity indicates that the generated outputs predominantly feature confidence phrases rather than hedging phrases. It reveals the overall characteristics of the generated outputs of the solved cases. Per case review of the confidence phrases showed the following averages: “evidence” appeared 1456 times (mean 6.330 per case), “final” appeared 1371 times (mean 5.961 per case), “Most disease-casing variants” were found 699 times (mean 3.039 per case),“clearly” appeared 236 times (mean 1.026 per case), “significantly” appeared 70 times (mean 0.304 per case), “established” appeared 36 times (mean 0.157 per case), subsequently, the terms “strongly”, “confirmed”, “robustly”, “conclusively”, and “certainly” were also observed.

Supplementary Chapter (Example from the parsed Genereviews dataset):

An example of a GeneReviews dataset generated through our approach employing the DeepSeek model: {gene": "*BRCA1*", "phenotype": "Hereditary breast and ovarian cancer", "priority": "Rank 1", "key_findings": "*BRCA1*-associated HBOC is characterized by increased risk for female and male breast cancer, ovarian cancer (including fallopian tube and primary peritoneal cancers), and to a lesser extent other cancers such as prostate cancer, pancreatic cancer, and melanoma. Risk varies depending on *BRCA1* vs *BRCA2* pathogenic variant.", "age": "Breast cancer diagnosed at or before age 50 years; ovarian cancer risk begins earlier in *BRCA1* carriers", "sex": "Both females and males", "inheritance_pattern": "Autosomal dominant", "mutation_type": "Heterozygous germline pathogenic variants", "severity": "High-grade tumors, triple-negative breast cancer phenotype", "clinical_manifestation": "Medullary histopathology, higher histologic grade, estrogen receptor negative, progesterone receptor negative", "comorbidities": "Multiple primary breast cancers, contralateral breast cancer"}.

### References Appendix:

11.	*GeneReviews®*. (University of Washington, Seattle, Seattle (WA), 1993).
